# Mesenchymal stromal cells for COVID-19: A living systematic review protocol

**DOI:** 10.1101/2020.04.13.20064162

**Authors:** Gabriel Rada, Javiera Corbalán, Patricio Rojas, COVID-19 L·OVE Working Group

## Abstract

**Objective:** To determine the impact of mesenchymal stromal cells outcomes important to patients with COVID-19.

**Design:** This is the protocol of a living systematic review.

**Data sources:** We will conduct searches in PubMed/Medline, Embase, Cochrane Central Register of Controlled Trials (CENTRAL), grey literature and in a centralised repository in L·OVE (Living OVerview of Evidence). L·OVE is a platform that maps PICO questions to evidence from Epistemonikos database. In response to the COVID-19 emergency, L·OVE was adapted to expand the range of evidence it covers and customised to group all COVID-19 evidence in one place. The search will cover the period until the day before submission to a journal.

**Eligibility criteria for selecting studies and methods:** We adapted an already published common protocol for multiple parallel systematic reviews to the specificities of this question.

We will include randomised trials evaluating the effect of mesenchymal stromal cells versus placebo or no treatment in patients with COVID-19. Randomised trials evaluating other coronavirus infections, such as MERS-CoV and SARS-CoV, and non-randomised studies in COVID-19 will be searched in case we find no direct evidence from randomised trials, or if the direct evidence provides low- or very low-certainty for critical outcomes.

Two reviewers will independently screen each study for eligibility, extract data, and assess the risk of bias. We will pool the results using meta-analysis and will apply the GRADE system to assess the certainty of the evidence for each outcome.

A living, web-based version of this review will be openly available during the COVID-19 pandemic. We will resubmit it every time the conclusions change or whenever there are substantial updates.

**Ethics and dissemination:** No ethics approval is considered necessary. The results of this review will be widely disseminated via peer-reviewed publications, social networks and traditional media.

**PROSPERO Registration:** Submitted to PROSPERO (awaiting ID allocation).

## INTRODUCTION

COVID-19 is an infection caused by the SARS-CoV-2 coronavirus [1]. It was first identified in Wuhan, China, on December 31, 2019 [2]; three months later, almost half a million cases of contagion had been identified across 197 countries [3]. On March 11, 2020, WHO characterised the COVID-19 outbreak as a pandemic [1].

While the majority of cases result in mild symptoms, some might progress to pneumonia, acute respiratory distress syndrome and death [4],[5],[6]. The case fatality rate reported across countries, settings and age groups is highly variable, but it ranges from about 0.5% to 10% [7]. In hospitalised patients it has been reported to be higher than 10% in some centres [8].

Mesenchymal stromal cells, previously known as mesenchymal stem cells [9], exhibit the capacity of homing to sites of injury and inflammation and exert antiinflammatory and immunomodulatory effects [10]. These cells can affect the status of T cells and skew them towards a regulatory phenotype, they also interact with B cells by inhibiting B cell responses [11]. Moreover, high susceptibility of mesenchymal stromal cells to viral infections in vitro could impair the clinical efficacy during an active viral infection [12].

Initial studies in COVID-19 patients suggest mesenchymal stromal cells would be able to reduce inflammation [13]. Several findings support this hypothesis, such as the expression of antiinflammatory and trophic factors, and the decrease of C-reactive protein, tumor necrosis factor-alpha and cytokine-secreting immune cells [14]. Additionally, the mesenchymal stromal cells infused to patients with COVID-19 were not infected by SARS-CoV-2 [14]. In consequence, it is expected that these cells could attenuate the overactivation of the immune system and support repair by modulating the lung microenvironment after SARS-CoV-2 infection [15].

Nonetheless, the record for the research about mesenchymal stromal cells to treat a vast array of diseases is not spectacular. Sound evidence of safety and efficacy is still lacking, despite hundreds of studies with the claim that they constitute revolutionary treatments [16].

Using innovative and agile processes, taking advantage of technological tools, and resorting to the collective effort of several research groups, this living systematic review aims to provide a timely, rigorous and continuously updated summary of the evidence available on the role of mesenchymal stromal cells in the treatment of patients with COVID-19.

## METHODS

This manuscript complies with the ‘Preferred Reporting Items for Systematic reviews and Meta-Analyses’ (PRISMA) guidelines for reporting systematic reviews and meta-analyses [17].

A protocol stating the shared objectives and methodology of multiple evidence syntheses (systematic reviews and overviews of systematic reviews) to be conducted in parallel for different questions relevant to COVID-19 was published elsewhere [18]. This protocol was adapted to the specificities of the question assessed in this review and submitted to PROSPERO (awaiting ID allocation).

## Search strategies

### Electronic searches

Our literature search was devised by the team maintaining the L·OVE platform (https://app.iloveevidence.com), using the following approach:

1. Identification of terms relevant to the population and intervention components of the search strategy, using Word2vec technology [19] to the corpus of documents available in Epistemonikos Database.
2. Discussion of terms with content and methods experts to identify relevant, irrelevant and missing terms.
3. Creation of a sensitive boolean strategy encompassing all the relevant terms
4. Iterative analysis of articles missed by the boolean strategy, and refinement of the strategy accordingly.

Our main search source will be Epistemonikos database (https://www.epistemonikos.org), a comprehensive database of systematic reviews and other types of evidence [20]. We supplemented it with articles from multiple sources relevant to COVID-19 [21].

In sum, Epistemonikos Database acts as a central repository. Only articles fulfilling Epistemonikos criteria are visible by users. The remaining articles are only accessible for members of COVID-19 L·OVE Working Group.

We will conduct additional searches using highly sensitive searches in PubMed/MEDLINE, the Cochrane Central Register of Controlled Trials (CENTRAL), Embase and the WHO International Clinical Trials Registry Platform.

The searches will cover from the inception date of each database until the day before submission. No study design, publication status or language restriction will be applied to the searches in Epistemonikos or the additional searches.

The following strategy will be used to search in Epistemonikos Database. We will adapt it to the syntax of other databases.

((coronavir* OR coronovirus* OR “corona virus” OR “virus corona” OR “corono virus” OR “virus corono” OR hcov* OR “covid-19” OR covid19* OR “covid 19” OR “2019-nCoV” OR cv19* OR “cv-19” OR “cv 19” OR “n-cov” OR ncov* OR “sars-cov-2” OR “sars-cov2” OR “SARS-Coronavirus-2” OR “SARS-Coronavirus2” OR (wuhan* AND (virus OR viruses OR viral)) OR (covid* AND (virus OR viruses OR viral)) OR “sars-cov” OR “sars cov” OR “sars-coronavirus” OR “severe acute respiratory syndrome” OR “mers-cov” OR “mers cov” OR “middle east respiratory syndrome” OR “middle-east respiratory syndrome” OR “covid-19-related” OR “SARS-CoV-2-related” OR “SARS-CoV2-related” OR “2019-nCoV-related” OR “cv-19-related” OR “n-cov-related”)) AND (MSC OR MSCs OR HMSC* OR stemstromal* OR stromalstem* OR nestcell* OR ((mesenchymal* OR “tissue-derived” OR “derived-mesenchymal”) AND (stromal* OR stem OR multipotent* OR progenitor*)) OR (medicinal* AND signalling* AND (cell OR cells)) OR (stromal* AND (stem OR multipotent*)) OR (“tissue-derived” AND mesenchymal*))

### Other sources

In order to identify articles that might have been missed in the electronic searches, we will do the following:

1. Screen the reference lists of other systematic reviews and evaluate in full text all the articles they include.
2. Scan the reference lists of selected guidelines, narrative reviews and other documents.
3. Conduct cross-citation search in Google Scholar and Microsoft Academic, using each included study as the index reference.
4. Review websites from pharmaceutical companies producing drugs claimed as effective for COVID-19, websites or databases of major regulatory agencies, and other websites specialised in COVID-19.
5. Email the contact authors of all the included studies to ask for additional publications or data on their studies, and for other studies in the topic.
6. Review the reference list of each included study.

## Eligibility criteria

### Types of studies

We will preferentially include randomised trials. However, information from non-randomised studies will be used if there is no direct evidence from randomised trials or the certainty of evidence for the critical outcomes resulting from the randomised trials is graded as low- or very low, and the certainty provided by the non-randomised evidence grades higher than the one provided by the randomised evidence [22].

We will exclude studies evaluating the effects on animal models or in vitro conditions.

### Types of participants

We will include trials assessing participants with COVID-19, as defined by the authors of the trials. Whenever we find substantial clinical heterogeneity on how the condition was defined, we will explore it using a sensitivity analysis.

In case we find no direct evidence from randomised trials, or if the evidence from randomised trials provides low- or very low-certainty evidence for critical outcomes, we will include information from randomised trials evaluating mesenchymal stromal cells in other coronavirus infections, such as MERS-CoV or SARS-CoV infections [22].

### Type of interventions

The interventions of interest are mesenchymal stromal cells obtained from any tissue. We will not restrict our criteria to any dosage, duration, timing or route of administration.

The comparison of interest will be placebo (mesenchymal stromal cells plus optimal treatment versus placebo plus optimal treatment) or no treatment (mesenchymal stromal cells plus optimal treatment versus optimal treatment).

Trials assessing mesenchymal stromal cells plus other interventions will be eligible if the cointerventions are identical in both intervention and comparison groups.

Trials evaluating mesenchymal stromal cells in combination with other active interventions versus placebo or no treatment will be also included.

### Type of outcomes

We will not use the outcomes as an inclusion criteria during the selection process. Any article meeting all the criteria except for the outcome criterion will be preliminarily included and assessed in full text.

We used the core outcome set COS-COVID [23], the existing guidelines and reviews and the judgement of the authors of this review as an input for selecting the primary and secondary outcomes, as well as to decide upon inclusion. The review team will revise this list of outcomes, in order to incorporate ongoing efforts to define Core Outcomes Sets (e.g. COVID-19 Core Outcomes [24].

#### Primary outcome

- All-cause mortality

#### Secondary outcomes

- Mechanical ventilation
- Extracorporeal membrane oxygenation
- Length of hospital stay
- Respiratory failure
- Serious adverse events
- Time to SARS-CoV-2 RT-PCR negativity

#### Other outcomes

- Acute respiratory distress syndrome
- Total adverse events

Primary and secondary outcomes will be presented in the GRADE ‘Summary of Findings’ tables, and a table with all the outcomes will be presented as an appendix [25].

### Selection of studies

The results of the literature search in Epistemonikos database will be automatically incorporated into the [L·OVE platform](https://test.iloveevidence.com/loves/5e6fdb9669c00e4ac072701d) (automated retrieval), where they will be de-duplicated by an algorithm comparing unique identifiers (database ID, DOI, trial registry ID), and citation details (i.e. author names, journal, year of publication, volume, number, pages, article title and article abstract).

The additional searches will be uploaded to the screening software Collaboratron™ [26].

In both L·OVE platform and Collaboratron™, two researchers will independently screen the titles and abstracts yielded by the search against the inclusion criteria. We will obtain the full reports for all titles that appear to meet the inclusion criteria or require further analysis to decide about their inclusion.

We will record the reasons for excluding trials in any stage of the search and outline the study selection process in a PRISMA flow diagram adapted for the purpose of this project.

### Extraction and management of data

Using standardised forms, two reviewers will independently extract data from each included study. We will collect the following information: study design, setting, participant characteristics (including disease severity and age) and study eligibility criteria; details about the administered intervention and comparison, including source of cells, dose, duration and timing (i.e. time after diagnosis); the outcomes assessed and the time they were measured; the source of funding of the study and the conflicts of interest disclosed by the investigators; the risk of bias assessment for each individual study.

We will resolve disagreements by discussion, and one arbiter will adjudicate unresolved disagreements.

### Risk of bias assessment

The risk of bias for each randomised trial will be assessed by using the ‘risk of bias’ tool (RoB 2.0: a revised tool to assess risk of bias in randomised trials) [27]. We will consider the effect of assignment to the intervention for this review. Two reviewers will independently assess five domains of bias for each outcome result of all reported outcomes and time points. These five domains are: bias due to (1) the randomisation process, (2) deviations from intended interventions (effects of assignment to interventions at baseline), (3) missing outcome data, (4) measurement of the outcome, and (5) selection of reported results. Answers to signalling questions and collectively supporting information will lead to a domain-level judgement in the form of ‘Low risk of bias’, ‘Some concerns’, or ‘High risk of bias’. These domain-level judgements will inform an overall ‘risk of bias’ judgement for each result. Discrepancies between review authors will be resolved by discussion to reach consensus. If necessary, a third review author will be consulted to achieve a decision.

We will assess the risks of bias of other study designs with the ROBINS-I tool (ROBINS-I: Risk Of Bias In Non-randomised Studies of Interventions) [28]. We will assess the following domains: bias due to confounding, bias in selection of participants into the study, bias in classification of interventions, bias due to deviations from intended interventions (effect of assignment to intervention), bias due to missing data, bias in measurement of outcomes and bias in the selection of the reported result. We will judge each domain as low risk, moderate risk, serious risk, critical risk, or no information, and evaluate individual bias items as described in ROBINS-I guidance. We will not consider time-varying confounding, as these confounders are not relevant in this setting [28].

We will consider the following factors as baseline potential confounders:

- Age
- Comorbidities (e.g. cardiovascular disease, renal disease, eye disease, liver disease)
- Co-interventions
- Severity, as defined by the authors (i.e respiratory failure vs respiratory distress syndrome vs ICU requirement).

### Measures of treatment effect

For dichotomous outcomes, we will express the estimate of treatment effect of an intervention as risk ratios (RR) or odds ratios (OR) along with 95% confidence intervals (CI).

For continuous outcomes, we will use mean difference and standard deviation (SD) to summarise the data using a 95% CI. Whenever continuous outcomes are measured using different scales, the treatment effect will be expressed as a standardised mean difference (SMD) with 95% CI. When possible, we will multiply the SMD by a standard deviation that is representative from the pooled studies, for example, the SD from a well-known scale used by several of the studies included in the analysis on which the result is based. In cases where the minimally important difference (MID) is known, we will also present continuous outcomes as MID units or inform the results as the difference in the proportion of patients achieving a minimal important effect between intervention and control [29].

Then, these results will be displayed on the ‘Summary of Findings Table’ as mean difference [29].

### Strategy for data synthesis

If we include more than one trial we will conduct meta-analysis for studies clinically homogeneous using RevMan 5 [30], using the inverse variance method with random effects model. For any outcomes where data were insufficient to calculate an effect estimate, a narrative synthesis will be presented.

### Subgroup and sensitivity analysis

We will perform subgroup analysis according to the definition of severe COVID-19 infection (i.e respiratory failure vs respiratory distress syndrome vs ICU requirement). In case we identify significant differences between subgroups (test for interaction <0.05) we will report the results of individual subgroups separately.

We will perform sensitivity analysis excluding high risk of bias studies, and if non-randomised studies are used, excluding studies that did not report adjusted estimates. In cases where the primary analysis effect estimates and the sensitivity analysis effect estimates significantly differ, we will either present the low risk of bias — adjusted sensitivity analysis estimates — or present the primary analysis estimates but downgrading the certainty of the evidence because of risk of bias.

### Assessment of certainty of evidence

The certainty of the evidence for all outcomes will be judged using the Grading of Recommendations Assessment, Development and Evaluation working group methodology (GRADE Working Group) [31], across the domains of risk of bias, consistency, directness, precision and reporting bias. Certainty will be adjudicated as high, moderate, low or very low. For the main comparisons and outcomes, we will prepare Summary of Findings (SoF) tables [32],[29] and also interactive Summary of Findings tables (http://isof.epistemonikos.org/). A SoF table with all the comparisons and outcomes will be presented as an appendix.

### Living evidence synthesis

An artificial intelligence algorithm deployed in the [Coronavirus/COVID-19 topic of the L·OVE platform](https://app.iloveevidence.com/loves/5e6fdb9669c00e4ac072701d) will provide instant notification of articles with a high likelihood to be eligible. The authors will review them, will decide upon inclusion, and will update the [living web version](https://app.iloveevidence.com/living-reviews/5e7e748c53e53e78053563fd) of the review accordingly. We will consider resubmission to a journal if there is a change in the direction of the effect on the critical outcomes or a substantial modification to the certainty of the evidence.

This review is part of a larger project set up to produce multiple parallel systematic reviews relevant to COVID-19 [18].

## Data Availability

All data related to the project will be available. Epistemonikos Foundation will grant access to data.

## Acknowledgements

The members of the COVID-19 L·OVE Working Group and [Epistemonikos Foundation](https://www.epistemonikos.cl/) have made possible to build the systems and compile the information needed by this project. Epistemonikos is a collaborative effort, based on the ongoing volunteer work of over a thousand contributors since 2012.

## NOTES

### Roles and contributions

GR conceived the common protocol for all the reviews being conducted by the COVID-19 L·OVE Working Group. GR drafted the manuscript, and all other authors contributed to it. The corresponding author is the guarantor and declares that all authors meet authorship criteria and that no other authors meeting the criteria have been omitted.

The COVID-19 L·OVE Working Group was created by Epistemonikos and a number of expert teams in order to provide decision makers with the best evidence related to COVID-19. Up-to-date information about the group and its member organisations is available here: epistemonikos.cl/working-group

### PROSPERO registration

This protocol has been submitted (awaiting PROSPERO ID allocation).

### Ethics

As researchers will not access information that could lead to the identification of an individual participant, obtaining ethical approval was waived.

## REFERENCES

1. World Health Organization. Director-General’s remarks at the media briefing on 2019-nCoV on 11 February 2020. [Internet] World Health Organization; 2020 [Accessed 2020 April 12] Available from:https://www.who.int/dg/speeches/detail/who-director-general-s-remarks-at-the-media-briefing-on-2019-ncov-on-11-february-2020.

2. Hui DS, I Azhar E, Madani TA, et al. The continuing 2019-nCoV epidemic threat of novel coronaviruses to global health - The latest 2019 novel coronavirus outbreak in Wuhan, China. Int J Infect Dis. 2020 Feb;91:264-266. Available from: doi:10.1016/j.ijid.2020.01.009

3. Dong E, Du H, Gardner L. An interactive web-based dashboard to track COVID-19 in real time. Lancet Infect Dis. 2020 Feb 19 Available from: doi:10.1016/S1473-3099(20)30120-1

4. Guan WJ, Ni ZY, Hu Y, et al. Clinical Characteristics of Coronavirus Disease 2019 in China. N Engl J Med 2020. Available from: doi:10.1056/NEJMoa2002032

5. Tavakoli A, Vahdat K, Keshavarz M. Novel Coronavirus Disease 2019 (COVID-19): An Emerging Infectious Disease in the 21st Century. BPUMS. 2020;22(6):432-450. Available from: doi:10.29252/ismj.22.6.432

6. Li LQ, Huang T, Wang YQ, Wang ZP, Liang Y, Huang TB, et al. 2019 novel coronavirus patients’ clinical characteristics, discharge rate and fatality rate of meta-analysis. Journal of medical virology. 2020. Available from: doi:10.1002/jmv.25757

7. Global Covid-19 Case Fatality Rates [Internet] UK: Centre for Evidence-Based Medicine [Accessed 2020 April 12]. Available from: https://www.cebm.net/covid-19/global-covid-19-case-fatality-rates/

8. Rodriguez-Morales AJ, Cardona-Ospina JA, Gutiérrez-Ocampo E, et al. Clinical, laboratory and imaging features of COVID-19: A systematic review and meta-analysis. Travel medicine and infectious disease. 2020;101623.Available from: doi:10.1016/j.tmaid.2020.101623

9. Viswanathan S, Shi Y, Galipeau J, Krampera M, Leblanc K, Martin I, et al. Mesenchymal stem versus stromal cells: International Society for Cell & Gene Therapy (ISCT®) Mesenchymal Stromal Cell committee position statement on nomenclature. Cytotherapy. 2019 Oct;21(10):1019-1024. Available from: doi: 10.1016/j.jcyt.2019.08.002

10. Kallmeyer K, Pepper MS. Homing properties of mesenchymal stromal cells. Expert Opin Biol Ther. 2015 Apr;15(4):477-9. Available from: doi: 10.1517/14712598.2015.997204

11. Elgaz S, Kuçi Z, Kuçi S, Bönig H, Bader P. Clinical Use of Mesenchymal Stromal Cells in the Treatment of Acute Graft-versus-Host Disease. Transfus Med Hemother. 2019;46(1):27–34. Available from: doi:10.1159/000496809

12. Cheung MB, Sampayo-Escobar V, Green R, Moore ML, Mohapatra S, Mohapatra SS. Respiratory Syncytial Virus-Infected Mesenchymal Stem Cells Regulate Immunity via Interferon Beta and Indoleamine-2,3-Dioxygenase. PLoS One. 2016;11(10):e0163709. Published 2016 Oct 3. Available from: doi:10.1371/journal.pone.0163709

13. Shetty, Ashok K. Mesenchymal Stem Cell Infusion Shows Promise for Combating Coronavirus (COVID-19)-Induced Pneumonia. Aging Dis. 2020. Available from: doi: 10.14336/AD.2020.0301

14. Leng Z, Zhu R, Hou W, Feng Y, Yang Y, Han Q, et al. Transplantation of ACE2(-) Mesenchymal Stem Cells Improves the Outcome of Patients with COVID-19 Pneumonia. Aging Dis. 2020 Mar 9;11(2):216-228. Available from: doi: 10.14336/AD.2020.0228

15. Atluri S, Manchikanti L, Hirsch JA. Expanded Umbilical Cord Mesenchymal Stem Cells (UC-MSCs) as a Therapeutic Strategy in Managing Critically Ill COVID-19 Patients: The Case for Compassionate Use. Pain physician. 2020;23(2):E71-E83. Available from: https://www.painphysicianjournal.com/current/pdf?article=NzAyNA%3D%3D

16. Marks PW, Witten CM, Califf RM. Clarifying Stem-Cell Therapy’s Benefits and Risks. N Engl J Med. 2017 Mar 16;376(11):1007-1009.Available from: doi: 10.1056/NEJMp1613723

17. Moher D, Shamseer L, Clarke M, et al. Preferred reporting items for systematic review and meta-analysis protocols (PRISMA-P) 2015 statement. Syst Rev. 2015 Jan 1;4:1. Available from: doi:10.1186/2046-4053-4-1

18. Rada G, Verdugo-Paiva F, Ávila C, Morel-Marambio M, Bravo-Jeria R, Pesce F, et al; COVID-19 L·OVE Working Group. Evidence synthesis relevant to COVID-19: a protocol for multiple systematic reviews and overviews of systematic reviews. Medwave 2020;20(3):e7867.Available from: doi:10.5867/medwave.2020.03.7867

19. Github repository [Internet] [Accessed 2020 April 12] Available from: https://github.com/dperezrada/keywords2vec

20. Epistemonikos Database Methods [Internet] Santiago: Epistemonikos Foundation [Accessed 2020 April 12]Available from: https://www.epistemonikos.org/en/about_us/methods

21. Methods for the special L·OVE of Coronavirus infection [Internet] Santiago: Epistemonikos Foundation [Accessed 2020 April 12] Available from: https://app.iloveevidence.com/covid-19

22. Schünemann HJ, Cuello C, Akl EA, Mustafa RA, Meerpohl JJ, Thayer K, et al; GRADE Working Group. GRADE guidelines: 18. How ROBINS-I and other tools to assess risk of bias in nonrandomized studies should be used to rate the certainty of a body of evidence. J Clin Epidemiol. 2019 Jul;111:105-114. Available from: doi:10.1016/j.jclinepi.2018.01.012

23. Xinyao Jin, Bo Pang, Junhua Zhang, et al. Core Outcome Set for Clinical Trials on Coronavirus Disease 2019 (COS-COVID), Engineering, 2020. Available from: doi:10.1016/j.eng.2020.03.002

24. COVID-19 Core Outcomes [Internet]. [Accessed 2020 April 12] Available from: https://www.covid-19-cos.org/

25. Guyatt GH, Oxman AD, Santesso N, et al. GRADE guidelines: 12. Preparing summary of findings tables-binary outcomes. J Clin Epidemiol 2013 Feb;66(2):158-72.Available from: doi:10.1016/j.jclinepi.2012.01.012

26. Collaboratron [Software]. Santiago: Epistemonikos Foundation, 2017.

27. Sterne JAC, Savović J, Page MJ, Elbers RG, Blencowe NS, Boutron I, et al. RoB 2: a revised tool for assessing risk of bias in randomised trials. BMJ. 2019 Aug 28;366:4898. Available from: doi:10.1136/bmj.l4898

28. Sterne JA, Hernán MA, Reeves BC, Savović J, Berkman ND, Viswanathan M, et al. ROBINS-I: a tool for assessing risk of bias in non-randomised studies of interventions. BMJ. 2016 Oct 12;355:i4919. Available from: doi:10.1136/bmj.i4919

29. Guyatt GH, Thorlund K, Oxman AD, et al. GRADE guidelines: 13. Preparing summary of findings tables and evidence profiles-continuous outcomes. J Clin Epidemiol 2013 Feb;66(2):173-83. Available from: doi:10.1016/j.jclinepi.2012.08.001

30. Review Manager (RevMan) [Software]. Version 5.3.5 Copenhagen: The Nordic Cochrane Centre, The Cochrane Collaboration, 2014.

31. Guyatt GH, Oxman AD, Vist GE, Kunz R, Falck-Ytter Y, Alonso-Coello P, et al; GRADE Working Group. GRADE: an emerging consensus on rating quality of evidence and strength of recommendations. BMJ. 2008 Apr 26;336(7650):924-6. Available from: doi:10.1136/bmj.39489.470347.AD

32. Guyatt GH, Oxman AD, Santesso N, et al. GRADE guidelines: 12. Preparing summary of findings tables-binary outcomes. J Clin Epidemiol [Internet] 2013 Feb [Accessed March 26] ;66(2):158-72. Available from: doi:10.1016/j.jclinepi.2012.01.012

